# Gene expression profiling of peripheral blood and endometrial cancer risk factors: Systems epidemiology approach in the NOWAC Postgenome Cohort Study

**DOI:** 10.1101/2025.03.09.25323623

**Authors:** Oxana Gavriluk, Igor Snapkow, Jean-Christophe Thalabard, Lars Holden, Marit Holden, Hege M. Bøvelstad, Eiliv Lund

## Abstract

The increasing incidence of endometrial cancer (EC) requires an extensive search for novel preventive tools and early intervention approaches. However, the establishment of reliable predictive models is impossible without knowledge of genetic changes prior to diagnosis. In this work, we aimed to establish whether known EC risk factors influence peripheral blood gene expression in a prospective design.

First, we selected variables (parity status, lifetime number of years of menstruation, coffee consumption, BMI, age of menopause, use of oral contraceptives) that were shown to have an impact on EC risk in a big prospective cohort (165,000 women). Next, using BeadChip microarray technology, we tested the association between these variables and changes in gene expression profiles in blood in a nested case-control study (79 case-control pairs) of women from the NOWAC postgenome cohort. Lastly, we undertook a gene set enrichment analysis (GSEA).

At overall gene expression level, we found no difference between the EC cases and controls. The introduction of parity status into the statistical model, revealed changes in the expression of 1,379 genes in the controls, while we did not observe any expression changes in the cases. Twenty-seven genes were associated with BMI increase in the controls, whereas there was no association observed between changes in BMI and gene expression in women with EC. In GSEA, 2407 significantly enriched gene sets were attributed to a parity increase among cancer-free women.

In this study, we found that an increased number of parities have a life-long effect on the gene expression profile in the peripheral blood of women who never developed cancer, while neither multiparity nor elevated BMI changed the gene expression in women diagnosed with EC later in life.

## Introduction

Endometrial cancer (EC) is the second most common and second most lethal gynecological cancer worldwide, with 7798 new cases registered in Norway between 2014 and 2023 [1, 2]. While the incidence and mortality rates of several other cancers have plateaued or decreased in the last decade, the incidence of EC has been rising globally, with the highest increase found in countries that have undergone a drastic change in living standards and lifestyles [3, 4]. In particular, changes in reproductive factors (e.g. declines in parity), combined with an increase in obesity prevalence, might explain the rise in EC incidence associated with socioeconomic transition [5].

Both descriptive and analytic epidemiological approaches can help health officials appropriately target prevention and control activities, but only provide limited information on the biological processes underlying the cause-effect relationship between a risk factor and the disease. Technological advances in genomic profiling provide epidemiologists with the opportunity to integrate molecular data into etiologic studies in order to decipher the carcinogenic process.

Thus far, most studies have been focused on the development of tools to reclassify endometrial tumors according to their molecular features, and stratify patients according to the risk of metastases and recurrence [6–8]. We recently showed that an in-depth analysis of multi-tissue genomic profiles could be a crucial addition to the personalized assessment of an individual’s biology and health [9].

As a major defense and transport system, blood immune cells can adjust the expression of their genes in response to various environmental factors and pathological conditions. We previously highlighted several specific behavioral programs, such as metabolism or signaling, deregulated in the individual’s blood cells, which are associated with biological and/or pathological responses to a given condition in the general population (e.g. smoking) [10], cancer-related risk factors [11] and health status (e.g. breast cancer diagnosis) [12, 13].

In this study, we aim to investigate whether the associations between known EC risk factors and gene expression changes in the blood cells are differential between women who will develop EC and the controls. This will help provide insight into the biological processes underlying lifestyle/exposure EC risk factors that might explain the incidence of EC.

## Material and methods

### The NOWAC Study

The NOWAC Study is a national population-based cohort study that includes Norwegian women between the ages of 30-70 randomly drawn from the Norwegian Central Population Register [14]. Starting from 1991 and within 4-6 years intervals, these women filled in questionnaires with a focus on lifestyle and health. Of this original cohort of approximately 172,000 women, more than 48,000 women born between 1943 and 1957 provided blood samples between 2003 and 2006, and filled out an additional two-page questionnaire to constitute the NOWAC Postgenome Cohort [14]. PAXgene tubes (PreAnalytiXGmbH, Hembrechtikon, Switzerland) were used to prevent RNA degradation after blood sampling, and allowed genome–wide analyses of blood gene expression profiles. Through the linkage to the Cancer Registry of Norway and The Cause of Death Registry in Statistics Norway, we have identified 88 women from the NOWAC Postgenome Cohort who developed EC between the time of blood sampling and December 31, 2008 (the end of follow-up) (Figure 1). Of these 88 individuals, four were excluded, as blood samples were not received and stored at −80C within four days after blood collection. To ensure the same storage time and age between the cases and controls, the controls, who did not receive any cancer diagnosis, were drawn at random from the NOWAC Postgenome Cohort, but matched by the time of blood collection and birth year. Matched case-control pairs of blood samples (n=84 pairs) were sent to the Genomics Core Facility at the Norwegian University of Science and Technology (NTNU) for microarray gene expression profiling in January 2011.

**Figure 1.**
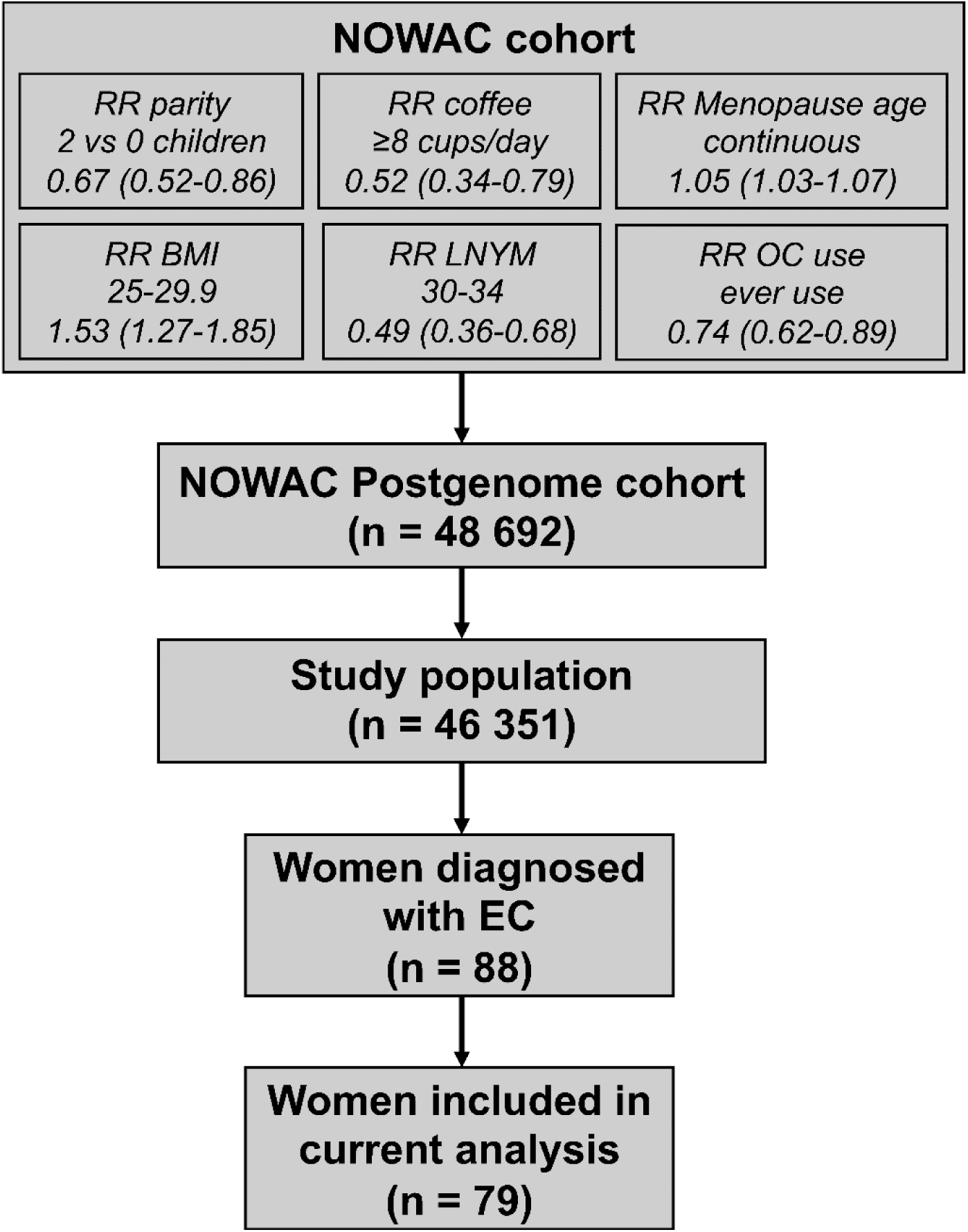
Study population. **Notes**: Relative risk estimates of endometrial cancer (EC) in the Norwegian Women and Cancer Study (NOWAC) were calculated using a multivariable model adjusted for body mass index (BMI), use of hormone replacement therapy (HRT), use of oral contraceptives (OC), smoking and alcohol consumption

### Laboratory procedures

In order to minimize the technical variability, each control sample was processed together with the matching case sample through RNA extraction, amplification and hybridization. Total RNA was isolated using the PAXgene Blood RNA Isolation Kit (Preanalytix, Qiagen, Hilden, Germany) following the manufacturer’s instructions. RNA quantity and purity were assessed by the NanoDrop ND1000 spectrophotometer (Thermo Scientific, Wilmington, Delaware, USA) and Agilent bioanalyzer (Agilent Technologies, Palo Alto, CA, USA) RNA amplification was performed in 96-well plates using 300 ng of total RNA and the Illumina® TotalPrep™-96 RNA Amplification Kit (Ambion Inc., Austin, TX, USA). Genome-wide RNA profiles were obtained using IlluminaHumanHT-12 chips version 4.

### Assessment of covariates and calculation of lifetime number of years of menstruation

Information on the covariates age at menarche, age at menopause, number of full-term pregnancies, duration of breastfeeding, height, weight, oral contraceptive use and smoking status was taken from NOWAC questionnaires (series from the years 2002-2005). Self-reported height and weight were used to calculate BMI in kg/m^2^. Parity and breastfeeding variables are generally reported to have a good validity in NOWAC Study [15]. The lifetime number of years of menstruation (LNYM) counts the number of years between age at menarche and age at menopause, minus the cumulative duration of full-term pregnancies (calculated as the number of full-term pregnancies, including live and stillbirths, times 0.75 years), duration of breastfeeding (calculated as the cumulative number of months of breastfeeding in all pregnancies) and duration of OC use [16].

### Outlier removal

The initial quality control made at the NTNU laboratory revealed two technical outliers (one 5S type degradation and one failed cRNA synthesis). These were removed along with their matching samples. Furthermore, one case was later found to have two cancer diagnoses, and was excluded along with its matching control. The data was carefully investigated to identify and remove technical outliers using the standard operating procedure for outlier removal described in [17]. In particular, probes related to genes in the human leukocyte antigen (HLA) systems were excluded (38 probes). These genes are known to be strongly expressed and have a high variance, which could affect multivariate analyses used in the outlier search. For individuals who were borderline outlier candidates, we excluded the ones with quality measures outside the range of the following thresholds: RIN value <7, 260/280 ratio <2, 260/230 ratio <1.7, and 50 < RNA < 500. Based on this outlier search, we identified two additional individuals as technical outliers, and excluded them along with their matching individual. In total, we investigated blood profiles from 79 women who developed EC and 79 age-matched controls (Figure 1).

### Microarray data preprocessing and normalization

Microarray data preprocessing and analysis were performed using R v3.3.1 (http://cran.r-project.org), in addition to tools from the Bioconductor project (http://www.bioconductor.org) adapted to our needs.

Expression profiles, including 47,248 probes, were adjusted for background noise using the negative control probes [18]. The data was further log2-transformed using a variance stabilizing technique [19], and finally normalized using quantile normalization. We retained probes present in at least 70% of the samples. If a gene was represented by more than one probe, the average expression of the probes was used as the expression value for the gene. The probes were translated to genes using the lumiHumanIDMapping database [20]. In total, the final data set included the expression for 7,104 unique genes. Finally, we computed the differences in log2 gene expression levels for each case-control pair.

### Statistical methods

We used linear models (Bioconductor R-package Limma [21]) to evaluate the significance of gene-wise expression differences between the cases and controls using the following intercept-only model:

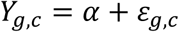

Where 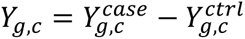 is the difference in the log2 gene expression level for case-control pair *c* and gene *g*, with ε_g,c_ normally distributed with zero expectation.

Using the same approach, we tested whether gene expression changes between the cases and controls were associated with EC risk factors (parity, LNYM, coffee, BMI, age of menopause, or OC use) using the following model:

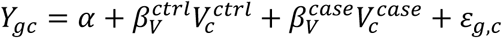

where 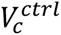 and 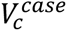 are the parity, coffee, LNYM, BMI, age of menopause or OC use variable for case-control pair *c* in the controls and cases, respectively.

Similarly, we applied a gene set approach to determine changes in gene signatures between the EC cases and controls associated with known risk factors. In this approach, the dependent variable is the difference in enrichment scores for case-control pair *c* and a defined gene set. The enrichment scores for eight collections (C1-C7 and H) of gene sets from the Molecular Signatures Database v6.1 (http://software.broadinstitute.org/gsea/index.jsp) [22] were obtained using the GSVA Bioconductor R package [23]. P-values were adjusted for multiple testing using the Benjamini-Hochberg procedure for controlling FDR [24].

Distributions of known EC risk factors were compared between the cases and matched controls using independent two-sided sample t-tests, Mann-Whitney U tests, and Chi square tests (R statistical package).

### Ethics Statement

The NOWAC study was approved by the Norwegian Data Inspectorate and the Regional Ethical Committee of North Norway (REK). The study was conducted in compliance with the Declaration of Helsinki, and all the participants gave their written informed consent. The linkages of the NOWAC database to national registries such as the Cancer Registry of Norway, The Cause of Death Registry, the National Population Register were approved by the Directorate of Health. The women were informed about these linkages. Furthermore, the collection and storing of human biological material was approved by the REK in accordance with the Norwegian Biobank Act. Women were informed in the letter of introduction that the blood samples would be used for gene expression analyses.

## Results

### Study population

In total, blood gene expression profiles were analyzed from 79 women diagnosed with EC after blood collection and 79 women matched by year of birth and time of blood sampling, who did not receive any cancer diagnosis within the same interval after blood collection. There was no significant difference in age, the onset of menarche and number of children between the two groups (Table 1). In both groups, approximately half of the women were premenopausal. In general, the cases had a later occurrence of menopause compared to the controls. Additionally, we observed a difference in LNYM, with a trend towards an increase in the cases. In our study, the cumulative breastfeeding duration was the lowest in the controls. This can be explained by a large spread in the reported time of breastfeeding by women with EC. There were more OC users among the cases. Our study population was overweight, in particular, 64.6% of women with an EC diagnosis had a BMI >25. The controls drank slightly more coffee.

**Table 1.**
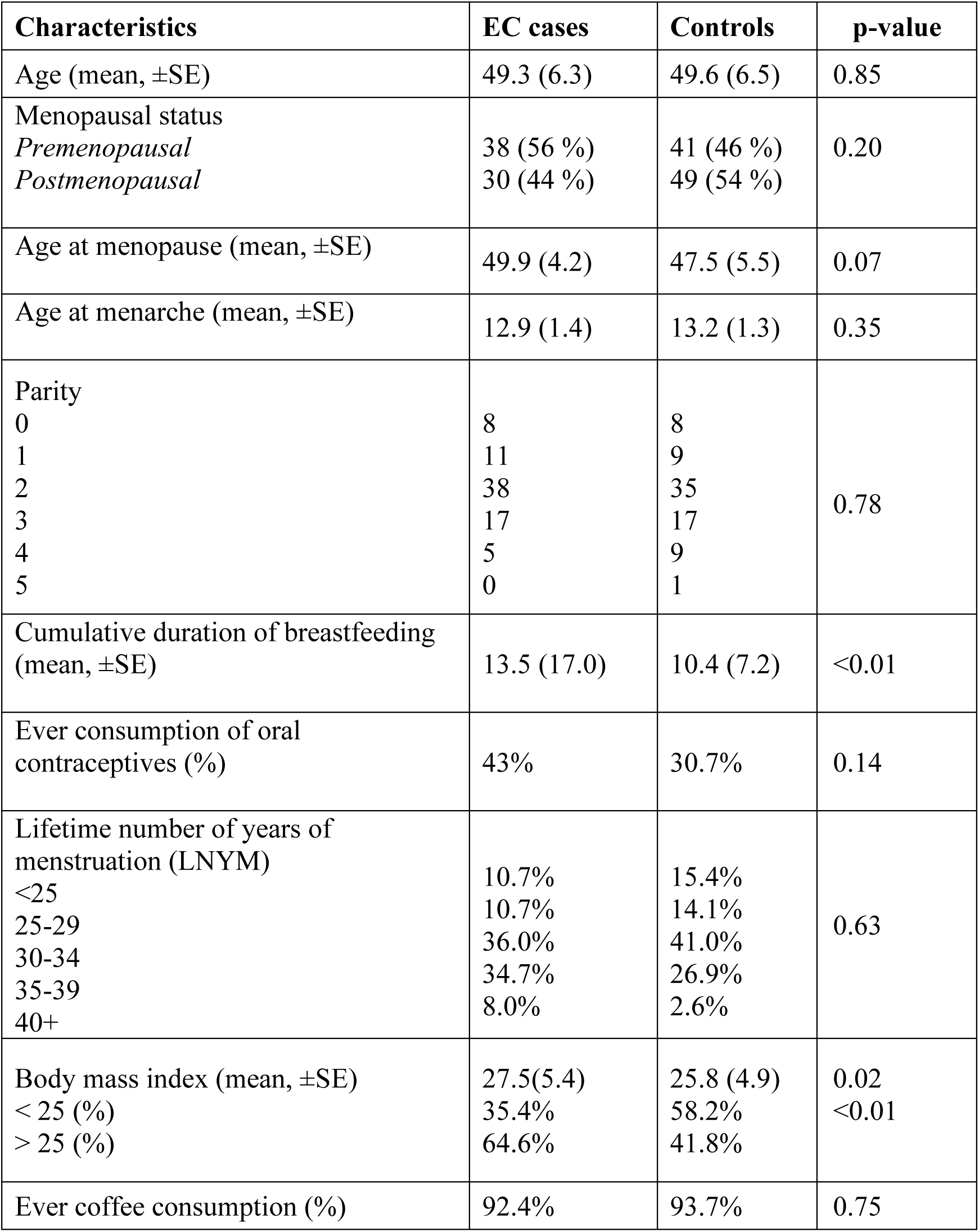
Baseline characteristics of the study (N = 79 case/control pairs)

### Differential blood gene expression profiles associated with EC diagnosis and risk factors in the cases and controls

After preprocessing, the study dataset included expression values for 7,104 genes. In the overall analysis, we were unable to identify any significant differences in gene expression profiles between the cases and controls. The same negative result was obtained when we performed a separate analysis for each year before diagnosis.

We then tested the hypothesis that the expression of some genes in the blood of either the cases or controls might be influenced by a set of variables modulating the risk of EC. Among the cases, there was no relationship between any of the variables used and log gene expression. In the controls, we observed no differentially expressed genes when using a model that included any of the following: coffee consumption, age of menopause, use of OC. Variations in BMI and the number of pregnancies had the strongest impact on the gene expression in the controls, where increasing parity was related to the expression differences of 1,379 genes (FDR 20%). A higher BMI altered the expression of 8, 17, and 27 genes at an FDR of 10%, 15%, and 20%, respectively. Of note, the major number of top 10 differentially expressed genes associated with both BMI and parity increase in the controls was downregulated (Table 2).

**Table 2.**
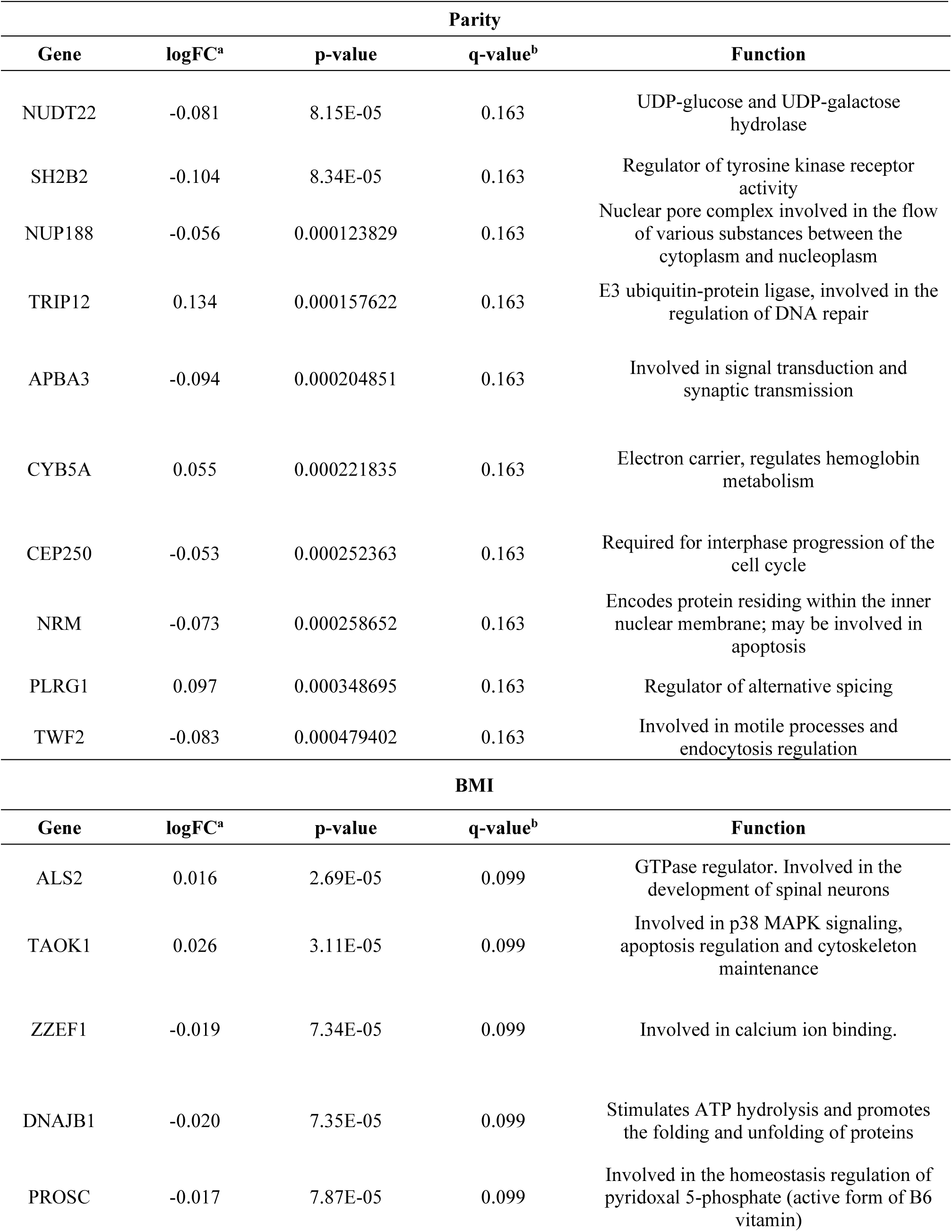
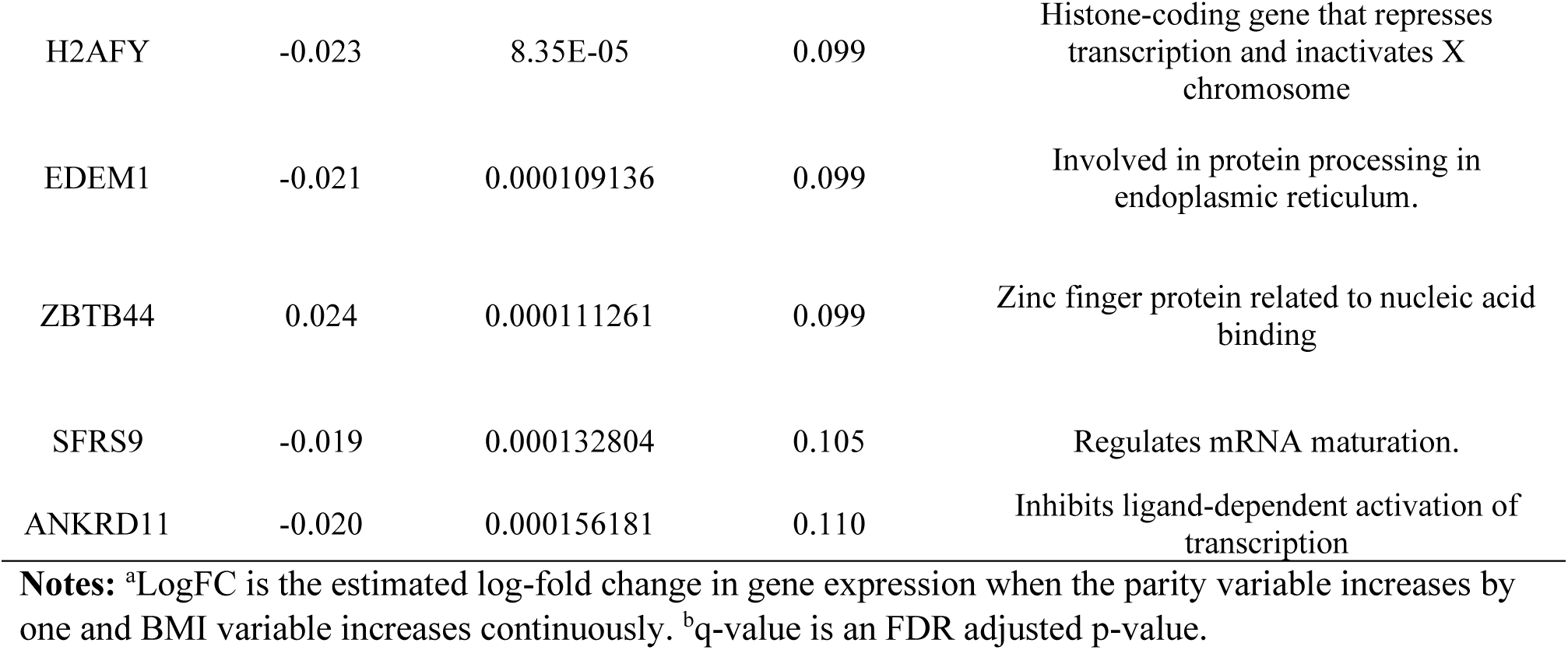
Top 10 differentially expressed genes associated with parity and BMI increase in the controls.

| Parity |  |  |  |  |
| --- | --- | --- | --- | --- |
| Gene | logFC<sup>a</sup> | p-value | q-value<sup>b</sup> | Function |
| NUDT22 | -0.081 | 8.15E-05 | 0.163 | UDP-glucose and UDP-galactose hydrolase |
| SH2B2 | -0.104 | 8.34E-05 | 0.163 | Regulator of tyrosine kinase receptor activity |
| NUP188 | -0.056 | 0.000123829 | 0.163 | Nuclear pore complex involved in the flow of various substances between the cytoplasm and nucleoplasm |
| TRIP12 | 0.134 | 0.000157622 | 0.163 | E3 ubiquitin-protein ligase, involved in the regulation of DNA repair |
| APBA3 | -0.094 | 0.000204851 | 0.163 | Involved in signal transduction and synaptic transmission |
| CYB5A | 0.055 | 0.000221835 | 0.163 | Electron carrier, regulates hemoglobin metabolism |
| CEP250 | -0.053 | 0.000252363 | 0.163 | Required for interphase progression of the cell cycle |
| NRM | -0.073 | 0.000258652 | 0.163 | Encodes protein residing within the inner nuclear membrane; may be involved in apoptosis |
| PLRG1 | 0.097 | 0.000348695 | 0.163 | Regulator of alternative splicing |
| TWF2 | -0.083 | 0.000479402 | 0.163 | Involved in motile processes and endocytosis regulation |
| BMI |  |  |  |  |
| Gene | logFC<sup>a</sup> | p-value | q-value<sup>b</sup> | Function |
| ALS2 | 0.016 | 2.69E-05 | 0.099 | GTPase regulator. Involved in the development of spinal neurons |
| TAOK1 | 0.026 | 3.11E-05 | 0.099 | Involved in p38 MAPK signaling, apoptosis regulation and cytoskeleton maintenance |
| ZZEF1 | -0.019 | 7.34E-05 | 0.099 | Involved in calcium ion binding. |
| DNAJB1 | -0.020 | 7.35E-05 | 0.099 | Stimulates ATP hydrolysis and promotes the folding and unfolding of proteins |
| PROSC | -0.017 | 7.87E-05 | 0.099 | Involved in the homeostasis regulation of pyridoxal 5-phosphate (active form of B6 vitamin) |
| H2AFY | -0.023 | 8.35E-05 | 0.099 | Histone-coding gene that represses transcription and inactivates X chromosome |
| EDEM1 | -0.021 | 0.000109136 | 0.099 | Involved in protein processing in endoplasmic reticulum. |
| ZBTB44 | 0.024 | 0.000111261 | 0.099 | Zinc finger protein related to nucleic acid binding |
| SFRS9 | -0.019 | 0.000132804 | 0.105 | Regulates mRNA maturation. |
| ANKRD11 | -0.020 | 0.000156181 | 0.110 | Inhibits ligand-dependent activation of transcription |

**Notes:** <sup>a</sup>LogFC is the estimated log-fold change in gene expression when the parity variable increases by one and BMI variable increases continuously. <sup>b</sup>q-value is an FDR adjusted p-value.

We also investigated gene overlap between the cases and controls according to the number of parities. A clear clustering among top differentially expressed genes associated with a changing number of parities either in the cases or controls was observed (Figure 2).

**Figure 2.**
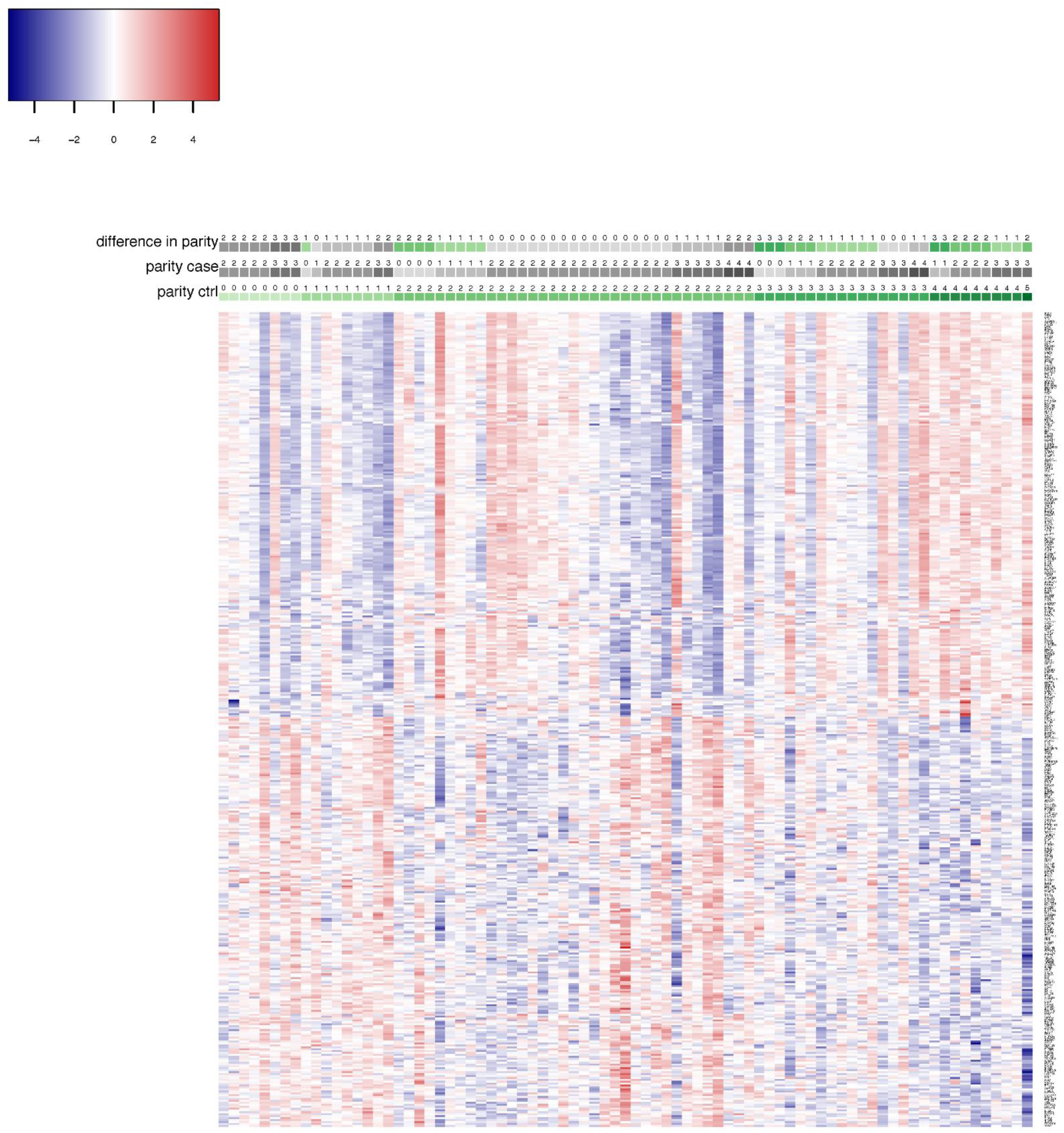
Top 100 genes overlap expression matrix for the cases and controls according to the parity status. Significant number of genes exhibits opposite expression pattern in the cases compared to the controls with increasing parity.

### Gene set enrichment analysis

For GSEA, we used all collections available at MSigDB (Molecular Signatures Database, http://software.broadinstitute.org/gsea/index.jsp) [22]. In total, for women diagnosed with EC, totally, we identified three significantly enriched gene sets (FDR 20%), of which one was parity-associated, one was enriched along with BMI increase and one was linked to the use of OC. Among the controls, GSEA revealed 2,415 enriched gene sets (FDR 20%), in which 2,407 were attributed to parity (Figure 3). Remarkably, the biggest part of these gene sets (786 at FDR 15% and 1,184 at FDR 20%) were from C7 collection (immunologic gene sets) (Figure 3).

**Figure 3.**
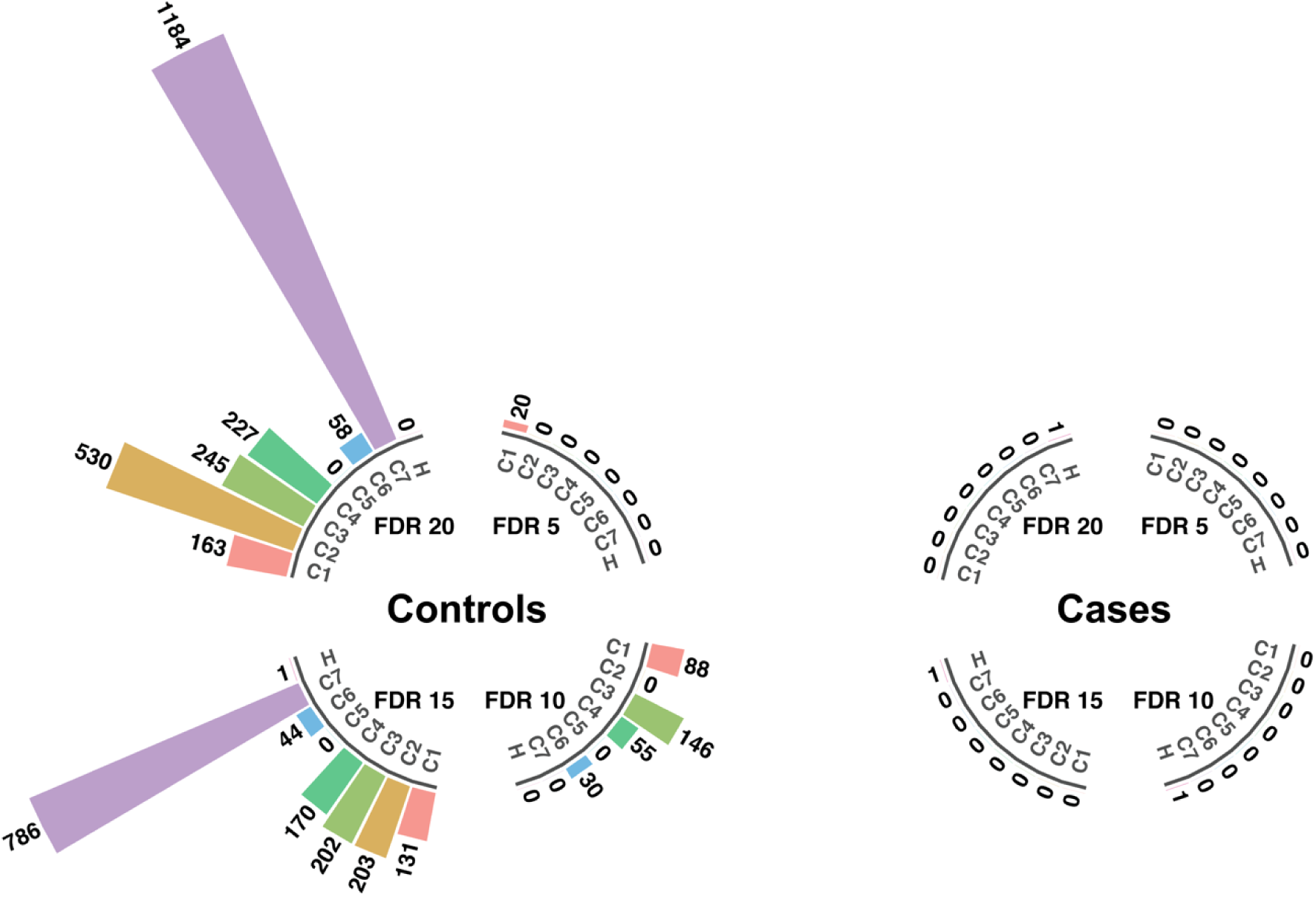
Number of significantly enriched gene sets in association with increasing parity. Circular barplot representing the influence of parity variable on gene set enrichment at different false discovery rate (FDR %) levels. Values on the top of the bars demonstrate the number of significant gene sets. C1, C2, C3, C4, C5, C6, C7, and H are respective gene set collections from MSigDB (described in Methods).

## Discussion

In a large prospective cohort, we studied the possibility to trace blood gene expression changes prior to EC diagnosis. We did not observe any significant differences in expression signatures between the cancer-free controls and women with EC when compared directly. Interestingly, BMI and parity, also known to be associated with EC incidence, were associated with significant changes in blood expression profiles in the controls, but not in the cases. To the best of our knowledge, this is the first study demonstrating pre-diagnostic blood gene expression differences between the EC cases and matched controls utilizing a systems epidemiology approach.

An association between a high BMI and an increased risk of EC development is well-established [25]. Moreover, overweight patients with EC have a higher risk of death, with a relative risk up to 6.25 in patients with BMI>40 compared to women of normal weight [26]. The main obesity-associated pathways contributing to EC development include augmented estrogen and estrogen metabolites synthesis, the presence of chronic inflammation and insulin resistance [25]. Unexpectedly, in our single gene analysis, we did not observe any changes in expression associated with a BMI increase in the EC cases. In turn, we observed a number of genes demonstrating a differential expression in the controls. This finding might be explained by the fact that the difference in BMI between the cases and controls in the study cohort was modest (Table 1). In GSEA, we identified three significantly enriched gene sets from C2 collection (one in the cases and two in the controls), and one gene set from H collection. Notably, the “HALLMARK_TGF_BETA_SIGNALING” gene set from H collection was enriched when the BMI of the controls increased. This finding is in line with other studies demonstrating the involvement of disturbed TGFβ signaling in EC development and progression [27, 28].

Not surprisingly, the major disparity on both single gene and gene sets levels in our study was connected to the number of pregnancies. There is a large body of evidence which shows the negative correlation between parity and the risk of EC [29–31]. However, a recent meta-analysis reports a nonlinear association between the number of children and RR [32]. Indeed, in the entire NOWAC cohort, we found a decrease in the EC incidence rate in women with one, two or three children compared to nulliparous (Figure 4). The elevated incidence rate of EC in women with four and more children is attributed to the low number of women with the high number of pregnancies in the cohort and, therefore, the limited sample size. Similar observations were reported by other studies [30]. A reduced time of estrogen exposure with increased parity is considered a major protective mechanism [33]. Furthermore, the shedding of the endometrium resulting in the mechanical elimination of potentially premalignant cells is well-described [32]. In the current study, we observed a significant enrichment of a large number of immunologic gene sets (C7 collection in MSigDB) in the controls with growing number of parities. Based on this finding, it is possible to assume that changes in the immune system associated with pregnancy may yet be another explanation of parity-dependent protection against EC. In addition, this protective effect cumulatively grows with every new child. Previously, our group published similar observations on parity and BC protection in a larger cohort [11]. Still, taking into account the complexity of gene sets data, limited sample size, and per se the explorative design of this study, it is impossible to provide a clear hypothesis on how the immune system changes in pregnancy contribute to EC protection. Therefore, further studies using both a laboratory and epidemiologic design, which address the long-term effects of immune processes on endometrial tumorigenesis, are warranted.

**Figure 4.**
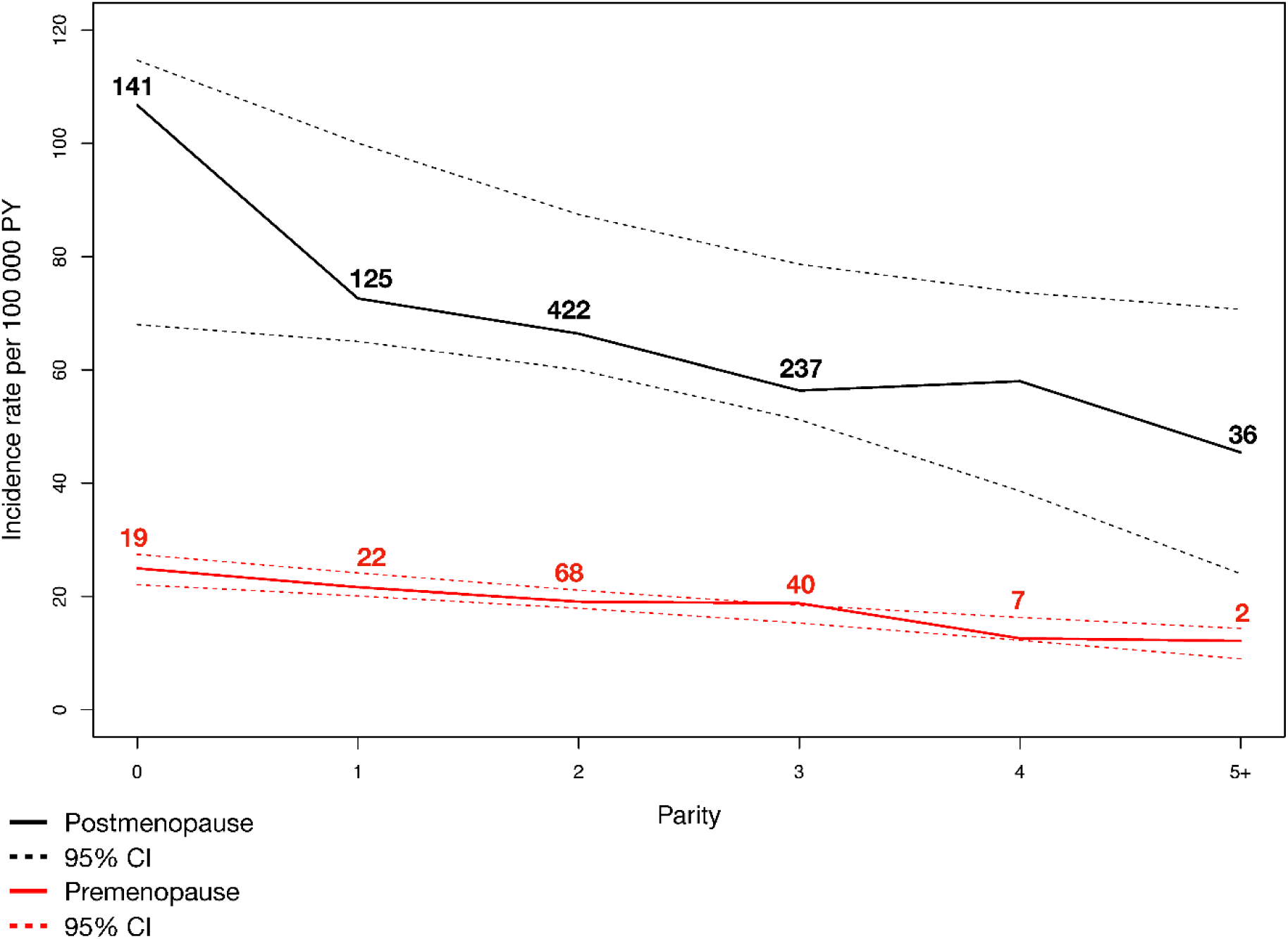
Endometrial cancer incidence rates according to parity and menopausal status for the NOWAC study 1991-2013 (n = 1,203)

In the cases, only the “HALLMARK_DNA_REPAIR” gene set was significantly associated with a high parity (FDR 10%). Alterations in DNA repair machinery are well-known to play a major role in EC carcinogenesis [34]. Thus, it is possible to hypothesize that mutations influencing DNA repair mechanisms could abrogate the protective effect of parity on EC and lead to cancer development, even in women who have given birth several times.

In current work, other factors with published evidences of involvement in EC development had a relatively weak impact on pre-diagnostic blood gene expression.

It has been demonstrated in NOWAC and by others that increased coffee consumption is inversely associated with EC risk [35, 36]. Here, we identified one significant gene set “HALLMARK_IL2_STAT5_SIGNALING” (FDR 20%) related to coffee drinking in the controls. Recently, Gotthardt and co-authors demonstrated that the maintenance of a stable STAT5 level is necessary for tumor surveillance by NK cells [37]. STAT5-depleted NKs were shown to promote tumor development. Hence, the impact of coffee compounds on the STAT5 metabolism in immune cells can be an additional biologic substrate of protective functions.

In OC users among cases, we revealed a significant enrichment of the “REACTOME_HYALURONAN_METABOLISM” gene set (FDR 20%). Interestingly, despite the low significance level, a second gene set from the top “REACTOME_HYALURONAN_UPTAKE_AND_DEGRADATION” was also related to hyaluronic acid metabolism. Elevated levels of hyaluronic acid in both tumor tissue and serum have been demonstrated to be involved in EC progression [38, 39]. Consequently, the monitoring of hyaluronan in the blood of women using OC might be a valuable tool in endometrial cancer screening.

Our findings of minor impact of increasing parity on peripheral blood gene expression in EC patients opposed to healthy women may suggest an impaired immune surveillance in EC.

To what extent circulating blood cells can reflect processes that occur in tumors is still an open question. Being an easily accessible tissue, blood may serve as an ideal tool for disease prognosis, monitoring and assessment of the treatment. In this work, we attempted to discover gene expression changes in circulating cells that can be identified long before a diagnosis of EC. It is important to emphasize that the findings reported here need further investigation, as most of the information available on the role of different genes in tumorigenesis is based on tissue studies and, therefore, cannot be entirely extrapolated to the blood cells.

The main strengths of the study include the prospective design and the population representativeness of the cohort, as well as complete information on cancer status, emigration and mortality obtained from national registries. The systems approach of testing epidemiologic data in the sub-cohort using gene expression profiles reduces the probability of false positive findings.

The relatively small sample size and the lack of a standardized algorithm for the gene expression analysis are limitations of this work. Moreover, the FDR levels we accepted were higher than recommended, although this can be justified by the relatively small sample size.

In conclusion, we identified a number of differences in gene set enrichment profiles between cancer-free women and women with a prior EC diagnosis in relation to the known risk factors for EC. We believe that this integrated analysis may provide a promising background for developing a new multilevel prediction model of EC risk at the population level. However, this approach should be further tested in a bigger sample size and in different populations.

## Data Availability

All data produced in the present study are available upon reasonable request to the authors

## Abbreviations

BMI: body mass index
CI: confidence interval
EC: endometrial cancer
FDR: false discovery rate
GSEA: gene set enrichment analysis
HRT: hormone replacement therapy
LNYM: lifetime number of years of menstruation
MHT: menopausal hormone therapy
NOWAC: The Norwegian Women and Cancer Study
OC: oral contraceptives

## Author’s contributions

OG, EL, VD and IS designed the study and interpreted the results. EL is a PI of the NOWAC Study. OG and IS constructed the tables, and drafted the manuscript. MH and LH carried out the statistical analyses, and contributed to drafting the manuscript. HMB participated in the initial statistical analysis, and critically revised the manuscript. VD and JCT contributed to the interpretation of the data, and to the drafting and revision of the manuscript. All authors read and approved the final manuscript.

